# Controlling the worldwide chaotic spreading of COVID-19 through vaccinations

**DOI:** 10.1101/2021.12.23.21268184

**Authors:** Hua Zheng, Aldo Bonasera

**Affiliations:** School of Physics and Information Technology, Shaanxi Normal University, Xi’an 710119, China; Cyclotron Institute, Texas A&M University, College Station, TX 77843-USA; Laboratori Nazionali del Sud, INFN, I-95123 Catania, Italy

**Keywords:** COVID-19, logistic map, vaccination

## Abstract

The striking differences and similarities between the ‘Spanish-flu’ of 1918 and the Coronavirus disease of 2019 (COVID-19) are analyzed. Progress in medicine and technology and in particular the availability of vaccines has decreased the death probability from about 2% for the Spanish-flu, to about 10^−4^ in the UK and 10^−3^ in Italy, USA, Canada, San Marino and other countries for COVID-19. The logistic map reproduces most features of the disease and may be of guidance for predictions and future steps to be taken in order to contrast the virus. We estimate 6.4 × 10^7^ deceases worldwide without the vaccines, this value decreases to 2.4 × 10^7^ with the current vaccination rate. In November 2021, the number of deceased worldwide was 5.1 × 10^6^. To reduce the fatalities further, it is imperative to increase the vaccination rate worldwide to at least 120 millions/day and the AstraZeneca vaccine due to its efficacity and cost is a possible route to accomplish this.

## 1 Introduction

The origins of the “Spanish-flu” of 1918 are unknown and so are the reasons why it stopped its deadly action in 1920, i.e., after a period of three years. Data from such disease are not complete as we would like, since science, medicine and technology were not sufficiently advanced. At that time, we were scientifically in better shape than in previous epidemics reported in history, but not enough to be able now to draw strict parallels with the Coronavirus disease first reported in 2019 (COVID-19). In 1918, we were at the end of World War I (WWI) and this probably contributed to the virus spreading in different continents so quickly. Furthermore, countries at war were not willing to share information about the disease in order to avoid advantages for the enemy. Spain, which did not participate in the conflict, openly reported about the high mortality rate of the disease in the country and this is the origin of the name Spanish-flu (S1918). Presently, there are no major wars but still some countries prefer not to make public all the relevant data. We know that there were many ‘waves’ in the period 1918-20, each more deadly than the previous one and it affected mostly young people. An explanation for this was that older people built their immune defenses from the pandemic of 1889-90 [1] but there is no definite proof about this. In any case it is a factor to be taken into account since ‘what does not kill you makes you stronger’. The 1889 pandemic was as serious as the S1918 but lasted a shorter time maybe due to the smaller flow of people travelling from different countries and continents. The many successive waves observed for the S1918 may be connected to seasonal variations or an evolution of the disease, i.e., different variants. The striking difference of the S1918 to the COVID-19 is the different age groups involved, most deaths in the latter case are older people even though new variants may change this unless we strengthen our immune defense through vaccines or survive to previous COVID-19 variants as in the 1889 case.

One hundred years ago we had little understanding of viruses, their spreading and how to contrast their deadly action. Masks and social distancing were adopted and some ‘homemade’ remedies with little if no efficacy. Thus, we can assume that the disease followed its course almost undisturbed. The method of transmission is through the vicinity of affected people to unaffected ones (thus the mask utility when no other defenses are available), where one person may affect n-persons with *n* ≥ 1 depending on the virus transmissibility. This can be expressed mathematically using chaotic maps and in particular we will discuss the logistic map:

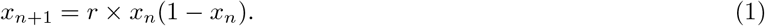

It is a simple iteration which, given an initial value 0 *< x*_0_ *<* 1, produces successive new values depending on the control parameter *r*. For *r* ≤ 4 the map is confined in the interval [0, 1], this is similar to the available phase-space in dynamical systems. For *r <* 3 the map gives two fixed points *x*_*f*_ which can be found assuming *x*_*n*+1_ = *x*_*n*_, they are *x*_*f*_ = 0 and *x*_*f*_ = 1 − 1*/r* [2]. For larger values of *r* a flip bifurcation occur, i.e., the map oscillates between two fixed values and this remains true for *r <* 3.449479, these values can be analytically recovered [2]. For larger values of *r*, the map oscillates among four fixed points (*r <* 3.544090) and so on, increasing the number of fixed points up to a critical value *r*_*c*_ = 3.569946 above which there are an infinite number of bifurcations and the map is fully chaotic [2, 3]. A powerful approach to quantify the ‘degree of chaoticity’ is to calculate an ensemble of *N* pairs of trajectories, separated initially by a very small distance *d*_0_. We introduce the mean distance between them at the iteration *n* as:

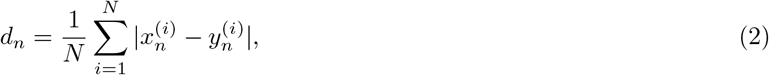

where

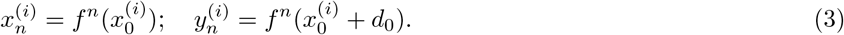

The initial starting points 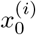 are chosen from a uniform distribution spanning the defining interval of the map. For fully chaotic maps the average distance *d*_*n*_ after *n*-iteration may be expressed by the relation

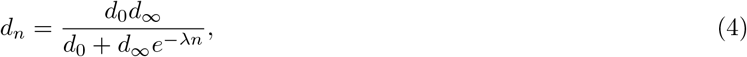

where the Lyapunov exponent *λ >* 0 indicates that nearby trajectories diverge to a finite (since the phase space is finite) value *d*_∞_(*<* 1). In the ergodic limit *r* = 4, *λ* = 2 which can be considered as the ‘highest chaoticity’ which can be reached. In the same limit [4] 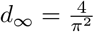, the largest average distance between nearby trajectories. Notice that even in the chaotic regime there are values of the control parameter where *λ <* 0, i.e., the map is not chaotic. We may argue that for the S1918 pandemic one of such values was accidentally hit and the epidemic disappeared suddenly. Notice that in such cases eq. (4) gives *d*_∞_ = *d*_0_ → 0 [4].

In refs. [5–8] we discussed the fact that the cumulative probability to be infected by COVID-19 (number of cases divided by number of tests) follows the same eq. (4) with the iteration *n* substituted by time *t*, thus if we know the Lyapunov exponents for the data and the map we can easily make a connection between time and iteration i.e.,

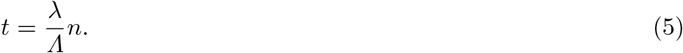

Under this assumption it follows that the maximum probability to become infected is 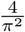. In 1918 the world population was about 2 billion which means that about 0.8 billions got infected by the S1918 in the period 1918-20. As we will discuss below about 2% of the infected died which gives 16 millions deaths, a number comparable to the casualties of WWI. Extending these estimates to 2020, the world population is about a factor 4 higher, which increases the estimates above by the same factor. However, in November 2021, 7.3 billion vaccine doses have been administrated which means that about 3.65 billion people have been fully vaccinated. Since the mortality rate is about 2% of the positive cases, then we expect the mortality rate (mostly non vaccinated people) to be given by (8 − 3.65) × 2% × 4*/π*^2^ = 35 millions. Also by the end of the year, people who received the first vaccine dose, will get the second thus reducing the maximum estimate to 18 millions. Optimistically 75% of the world population may be vaccinated (excluding children age less than 12 for whom vaccination is not possible yet, but more recently the use of some vaccines have been extended to children aged 5 or older). This gives a mortality rate 0.25 × 8 × 10^9^ × 2% × 4*/π*^2^ = 1.6 × 10^7^, currently, the number of recorded deaths is 5.1 million. Our estimate above assumes that the spreading is fully chaotic and as we will show below this is not the case. If we take Italy as an example, the average positive probability is about a factor 4 below the ergodic limit of the logistic map. These are in any case very important numbers thus the urgency to increase the number of vaccines worldwide, the faster we do the more we protect children and people with medical conditions and that cannot be vaccinated. We would like to stress that the extension of vaccinations to the youngest population may be necessary since there is some part of the population refusing to get vaccinated denying the clear success vaccines have obtained so far. But this is not enough especially if new variants will appear.

In the figure 1 (top panel) we plot the cumulative number of cases (positive and deceased) as function of time starting from January 1, 2020 for Italy (top-left panel) and the UK (top-right panel) [10, 11]. In order to directly compare the two countries we have divided the number of cases by their population (60,177,000 in Italy and 68,207,116 in the UK). The two cases are strikingly similar and we have observed no major differences to other countries, see also supplemental material. After the first rapid increase there is a plateau, which roughly starts at the end of the lockdowns and the beginning of the summer season. In the fall of 2020 the cases increase rapidly again forcing new lockdowns to slow them down and fortunately in December 2020 vaccinations started. The final increase in the UK is due to the Delta variant of the disease. The number of deceased is roughly a factor 50 smaller than the number of positives and no increase is seen at later times thanks to vaccinations. If we treat these data as representative of cases worldwide we can estimate a total number of positive to the virus in 790 millions (10% of the current world population) and a total number of deceased in 16 millions (0.2% of the current world population), in November 2021 those values are 253 millions and 5.1 millions deaths respectively. If this trend is followed, i.e., no increase in the number of vaccines and no other more deadly variant, we can estimate the maximum total duration of the disease in 5 years, two more than the S1918. These values are of course not acceptable and a worldwide effort is needed, especially vaccinations. We notice in passing that some countries, notably S. Korea and others, have adopted strict distancing, tracking and wearing mask measures, which successfully stopped the spreading, see supplemental material. Other countries have not been able to follow these examples because of unrest from part of the population thus leaving vaccinations and new medicines to contrast the spreading. Unfortunately, in the same countries there has been some resistance to vaccination as well but this may be overcome through compulsory vaccinations or the natural spreading of the virus from vaccinated to unvaccinated ones. In the latter case, the weak part of the population, which cannot be vaccinated, may pay a high price unwillingly. A fit to the top part of the figure 1 using eq. (4) gives about 7% and 0.2% total number of positives and deceased of the population respectively. These values are confirmed by many other countries, see supplements, with the notable exception of S. Korea reducing to 0.6% and 0.004% respectively. The approach and methods used by S. Korea and other Asiatic countries should be part of the civic study curricula of students starting in elementary schools worldwide to avoid the drama for the next pandemic, which could occur in the next 10 or 100 years.

**Fig. 1.**
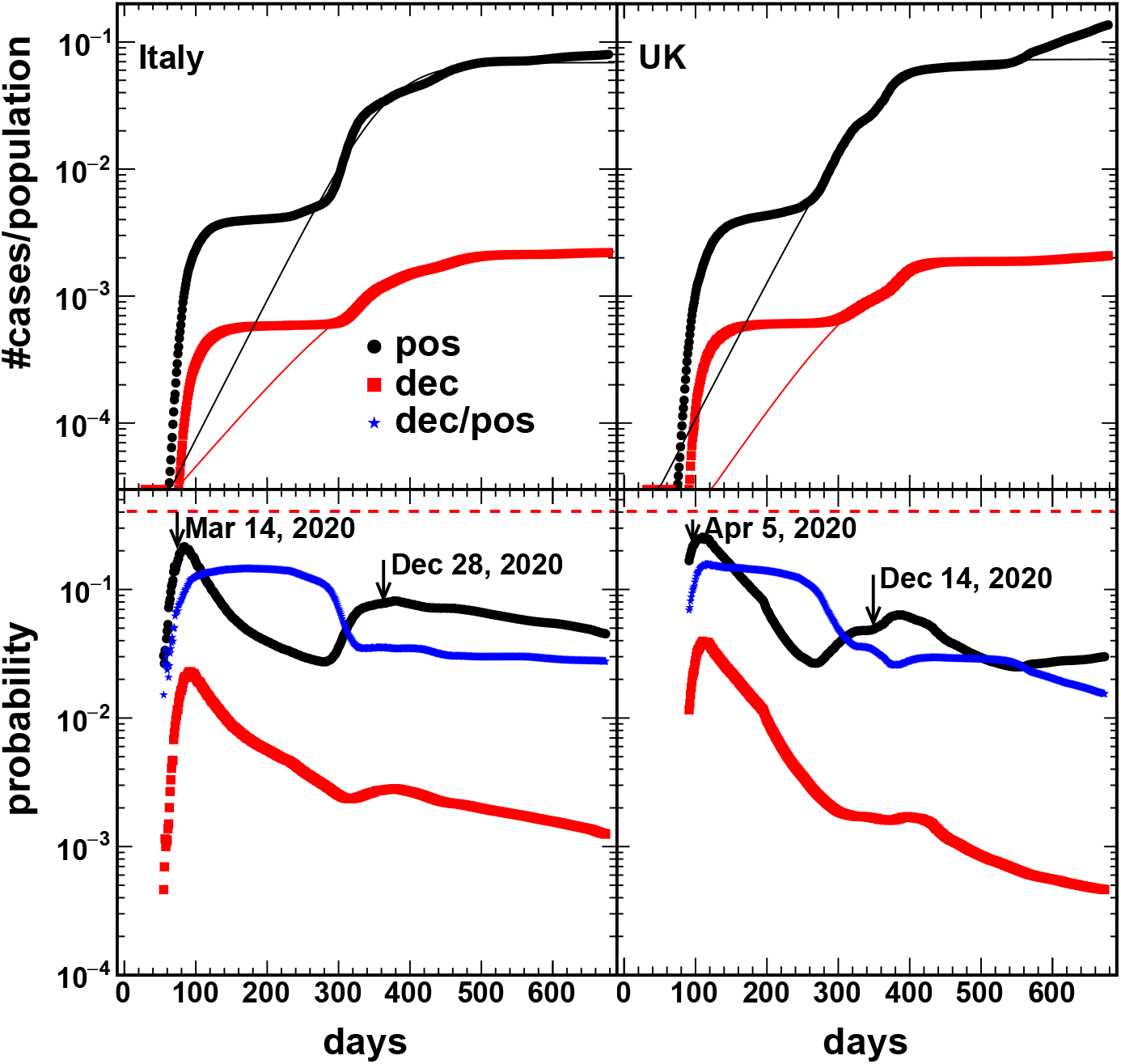
(Color online) Updated to November, 2021. (Top) The cumulative number of cases (positive and deceased) over population versus time for Italy and UK respectively. January 1, 2020 is the day 1; (Bottom) The corresponding probabilities as well as the ratio deceased over positive versus time. The arrows for March 14, 2020 and December 28, 2020 correspond to complete lockdown and start of vaccinations for Italy. Similarly, April 5, 2020 and December 14, 2020 correspond to the UK prime minister hospitalization for COVID-19 and start of vaccinations respectively.

The estimates above could be misleading because, especially at the very beginning of the disease, only part of the population was tested. Since the number of tests each day are known we can define probabilities as the number of cases divided the number of tests [5–8]. These probabilities are reported in the bottom panels of figure 1. The quantities reported in figure 1, top and bottom, would be the same if the total number of tests equals the countries’ population. The dashed lines in the bottom panels are given by the logistic map value 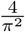 and in no cases it is reached apart Norway, see fig. S1, at the very beginning of the pandemic. In particular the rapid increase at the beginning reaches a maximum value on March 14, 2020 when the lockdown was imposed in Italy and April 5, 2020 when the UK prime minister was admitted at the hospital in serious conditions having contracted COVID-19. As we noticed before the deceased probability follows the same trend and it is about a factor 50 lower than the positives, shifted in time of about one week. Notice that the ‘population’ for the deceased is given by the positives, thus we can define another probability as the number of deceased divided by the number of positives. This quantity is plotted in the bottom part of figure 1 (blue symbols) as function of time and it is very close to the probability to be positive to COVID-19 as expected. In particular, after a first rapid increase at the beginning it plateaus at about 10% and jumps down to 2-3% again in December 2020 when the vaccination campaigns started, clearly demonstrating their efficacity. We stress that in the S1918 case, if the disease was completely out of control, then the number of deaths may be a factor of 5 higher than estimated above, i.e., of the order of 80 millions.

In order to compare to the logistic map, we plot in figure 2 the positive probabilities in two different periods for Italy (top panel) and UK (bottom panel). The logistic map follows rather well (red symbols) the data and the values of the Lyapunov and *d*_∞_ can be extracted for all cases. From ref. [4] we know that *d*_∞_ can be parametrized as

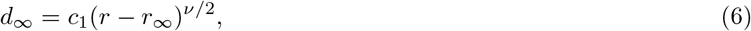

as function of *r* with 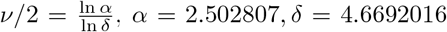 and *r*_∞_ = 3.569946. And similarly for the Lyapunov exponent

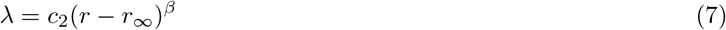

where 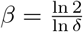. The values *c*_1_ and *c*_2_ can be fixed to their analytical values in the ergodic limit i.e., r=4.

**Fig. 2.**
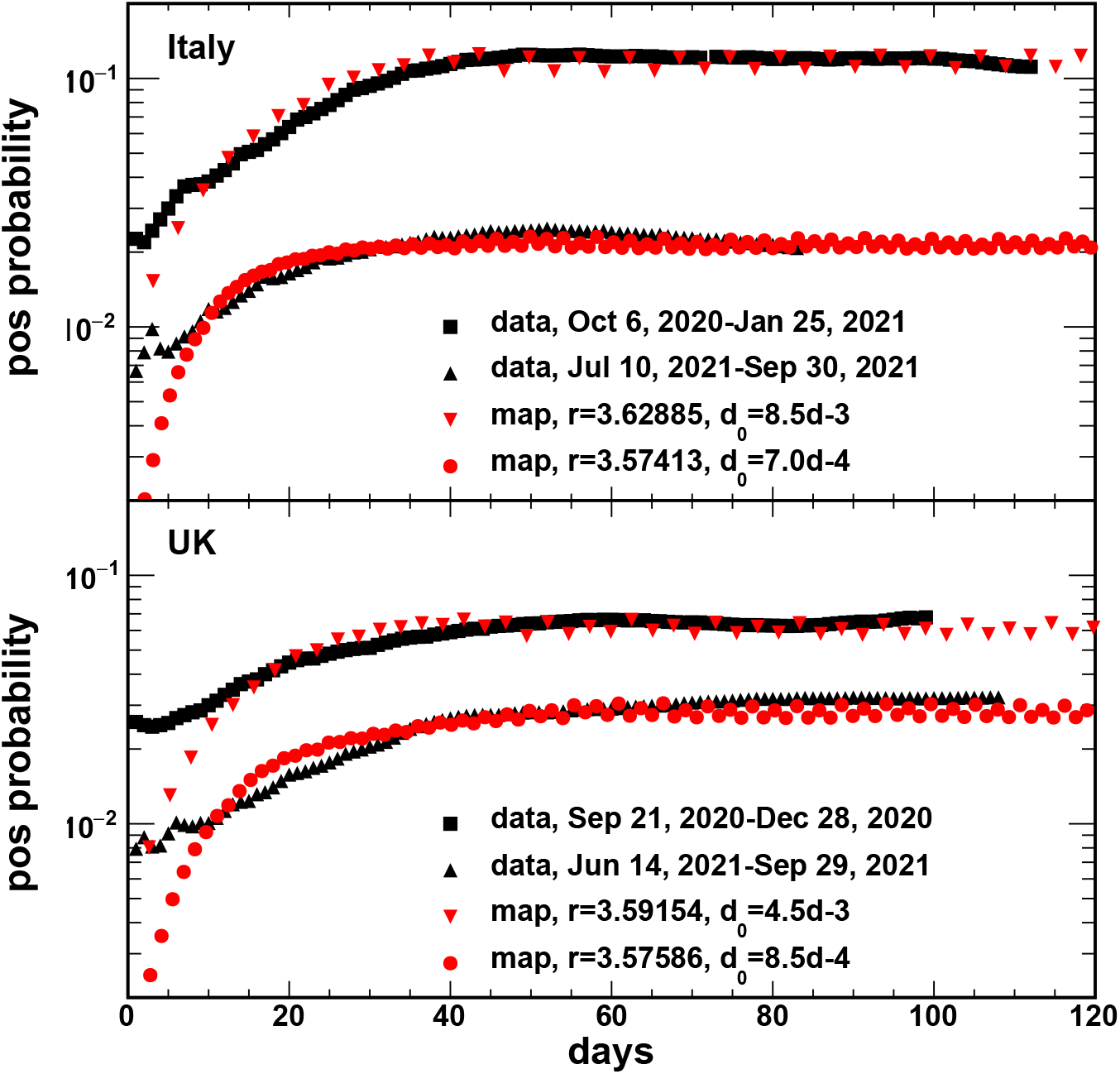
(Color online) The comparison between the logistic map results with the data in Italy (top panel) and UK (bottom panel) for the second wave in 2020 and the third wave in 2021.

From the number of positives we know the value of the asymptotic probability which plays the role of *d*_∞_ and it is the same as for the logistic map, thus we can extract the corresponding value of *r* from eq. (6) and, knowing *r* we obtain the Lyapunov exponent for the map eq. (7). From the ratio of the Lyapunov exponent for the map and the data we can make the transformation from the number of iterations to time, eq. (5). In table 1 we have collected some results for different countries. Large values for both, Lyapunov exponent and *d*_∞_, are obtained by Sweden which opted for herd immunization. Fortunately, the low population density and good health organization decreased the possible catastrophic effect of such a choice as we saw in the UK which were following the same strategy at the beginning of the pandemic [5–8]. In the last column of table 1 we report the value of *r* − *r*_∞_. If this quantiy becomes zero or negative then we have reached ‘herd-immunization’. As we see from the table, no country gets such values but it gets very close to it especially for the deaths probabilities which is the most important quantity. The combined action of vaccine and new effective medicines which reduces mortality may finally take us to herd-immunization. People may still get COVID-19 but will not die because of it!

**Table 1.**
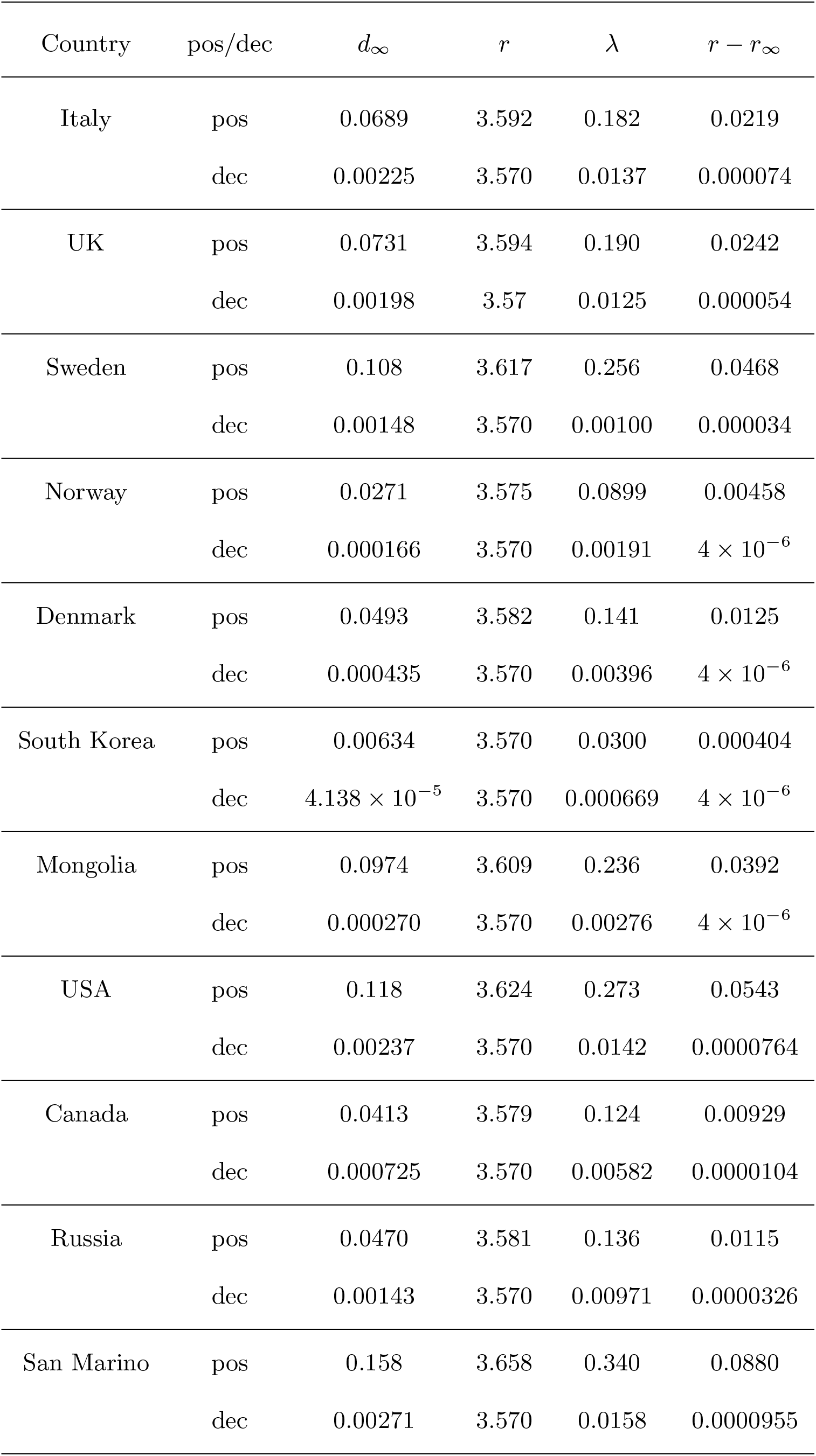
The *r* and *λ* calculated from Eqs. (6) and (7) for Fig. 1 and Fig. S1.

## 2 Controlling chaos

We have discussed in the introduction the connection between the control parameter *r* of the logistic map with *λ* and *d*_∞_. In order to have *r < r*_*c*_ we can decrease the value of *d*_∞_ through lockdowns, masks, tracing and vaccinations. In particular we can write:

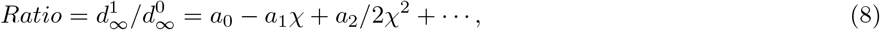

where, 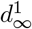 is the current value of the probability while 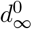 is the same quantity but exactly in the same period of the year before. *χ* is the controlling factor which we assume equal to

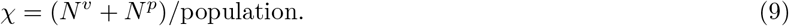

*N*^*v*^ and *N*^*p*^ are the number of vaccinated and positive to COVID-19 respectively, since *N*^*p*^ ≪ *N*^*v*^ it can be neglected for all practical estimations. The expansion in eq. (8) may need higher order terms because of lost of efficacity of the vaccine, of previous infections and/or new variants. Unvaccinated people contribute to eq. (9) if they get infected, thus another reason to get vaccinated as soon as possible. Also vaccinated people may be justified not wearing masks in presence of unvaccinated by choice, this may spread the virus faster when it is harmless to vaccinated and there are medicines which can save the life of the unvaccinated, keeping in mind that 2-3% of the positives will die, see figure 1. Future, more deadly variants may develop thus the importance to stop the epidemic as quickly as possible. In figure 3 we plot the probabilities as function of the vaccination ratio *χ*. The decrease is very encouraging but it maybe misleading since could be due to lockdowns or simply to a major awareness from the population. A more suitable quantity may be given by the ratio of probabilities or cases in two different periods, see eq. (8).

**Fig. 3.**
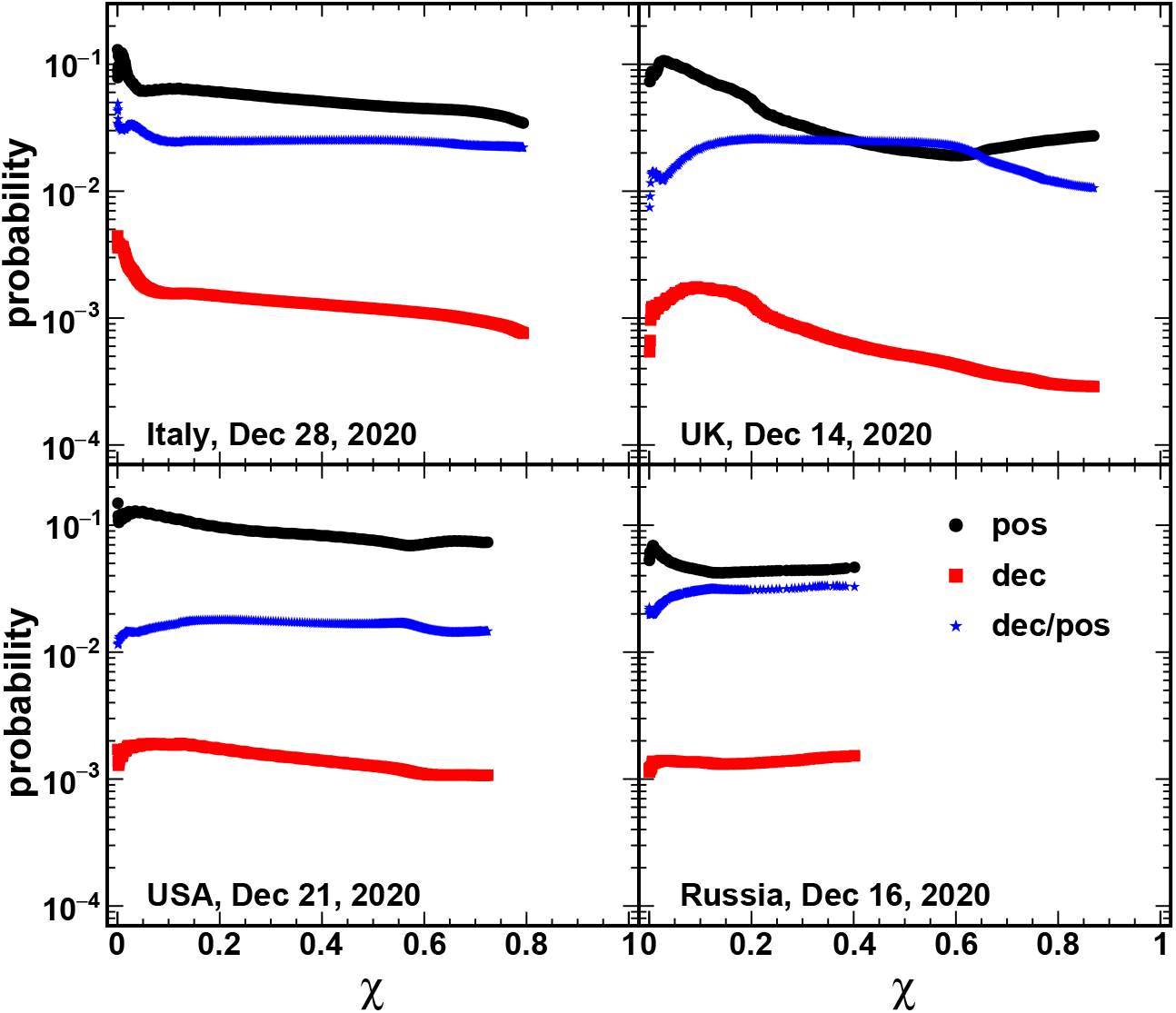
(Color online) The probabilities (positive, deceased and deceased/positive) versus the vaccination ratio *χ* for different countries as indicated. The date of the first day of vaccination has been indicated in the figure for each country.

From eq. (8) we can define the probability ratios for the year 2021 divided the same period in 2020. Since, in Italy for instance, test data was taken starting Feb 24, 2020 that is the first day we can build the ratios (for the UK, US and Russia are Mar 31, 2020, Mar 1, 2020 and Mar 4, 2020 respectively). This ratio is plotted in figure 4 as function of the parameter *χ* defined in eq. (9). The divergence for low vaccination rates is due to the low statistics at the very beginning of the pandemics when the virus was starting to diffuse among the people. From the expansion, eq. (8), we should get an initial decrease followed by higher order corrections that could be positive or negative. If 75% of the population is vaccinated, we expect the number of cases to decrease of a factor at least 4 as respect to the year before. The figure displays a fast decrease at the beginning of the vaccinations followed by a later increase due most probably to the Delta variant and the release of strong restrictions and lockdowns. The fact that the ratio for positives is about 1 for large *χ* values indicates that herd immunity cannot be reached. On the other hand the ratio for deceased is about 20% for the US and Italy and 4% for the UK indicating that vaccines are working in preventing deaths but with different efficacities. As we noticed above, the number of deaths should be compared to the number of positives and not to the number of tests, thus in the same figure we include this ratio with blue symbols. Now the results are comparable to the deceased probabilities as expected with the exception of Russia. The important lesson to take from this plot is that the vaccines are working and in particular the approach used in the UK seems to be the most effective. A striking difference is given by Russia, which follows the same trend as other countries but the ratios are larger than one and the deceased are above the positives, figure 4. This suggests that an important epidemic wave is underway in Russia more important than the previous year together with the resistance of people to get vaccinated. We had noticed in previous works [7] that in the year 2020 while the other countries were “under siege” from the virus, Russia was relatively free from it. The general behavior as function of vaccination confirms the quality of the vaccine used, in any case we can test this by analyzing cases from the Republic of San Marino (RSM), which adopted the same vaccine as Russia. The figure 4 shows an increase in the ratio with increasing vaccination rate. We have attempted fits using eq. (8) adding terms in the expansion up to 9th order. Those fits are quite bad showing strong oscillations thus confirming once more that we are nowhere near herd-immunity. This is not going in the direction of herd immunity but it could be an artifact due to the different number of tests in the different periods. To explore this feature we will discuss below the ratios of the number of cases in the two periods.

**Fig. 4.**
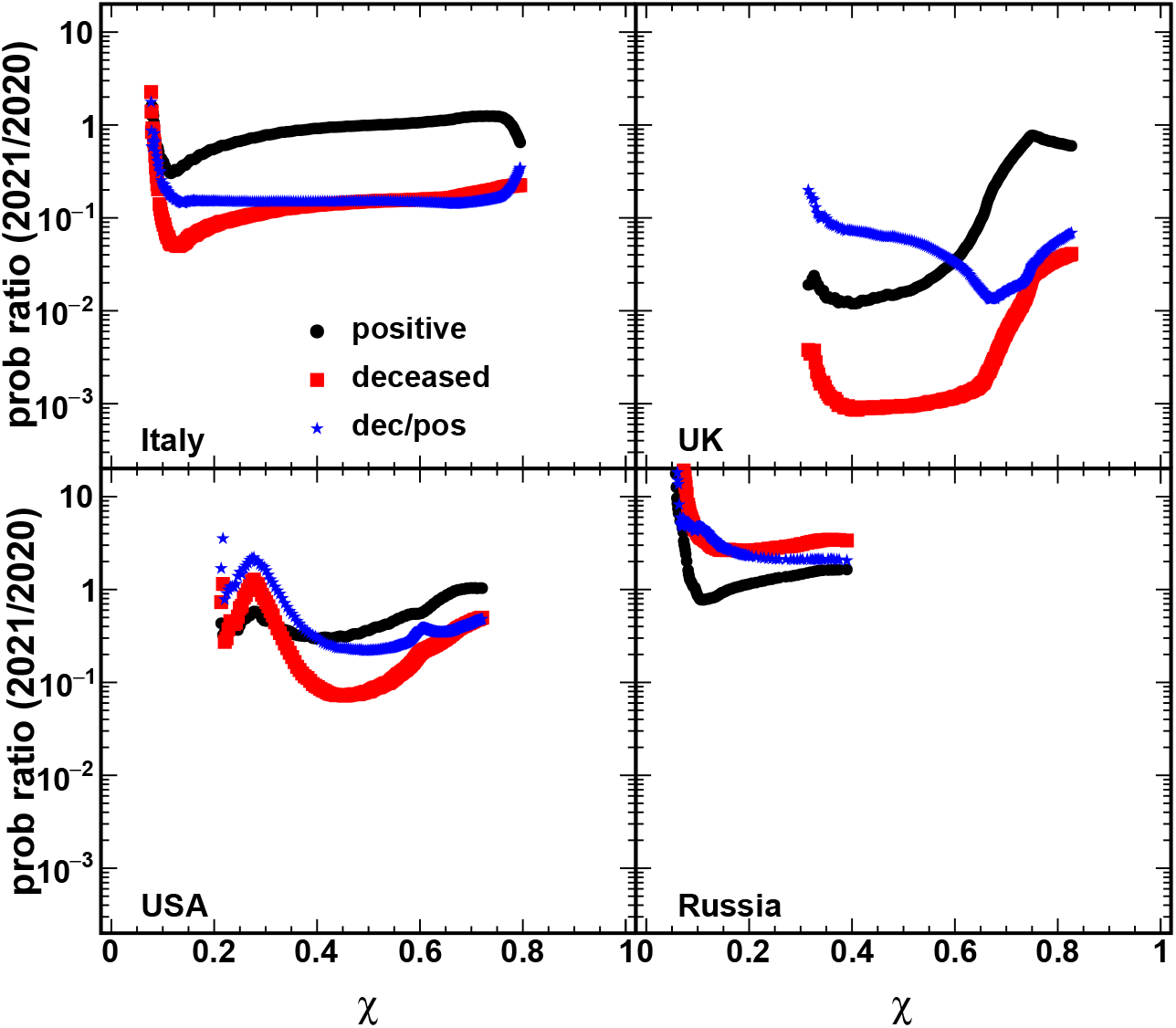
(Color online) The probability ratio for the same period in 2021 and 2020 versus the vaccination ratio *χ* for each country.

RSM does not provide the same data as for the cases in figure 4 but we can make the ratio of the number of deceased for the same period in the two years under study. In figure 5 (top panel) we plot the ratios as function of *χ* for different countries (top panel). We have added Canada as well since it has a large number of vaccinations. We also expect very many similarities between the UK and Canada, the weather, social organization etc., and we wanted to test if the different vaccines used in the two countries give different results. USA and Canada used the same vaccines (Moderna, Pfizer and Johnson&Johnson) and their behavior is very similar but slightly shifted. The RSM has lower statistics, thus higher fluctuations, but it plateaus at the lower values of the previous two countries, thus the Sputnik-V vaccine is as effective as the previous ones. The UK case, AstraZeneca vaccine used especially at the beginning of the vaccination campaign, gives much lower ratios thus much more effective than the other vaccines. These cases confirm the results discussed for figure 4, the different observables are consistent. Italy is slightly above the other countries but we believe it is due to the different ways of counting the deceased. A careful investigation should be performed. Other vaccines are used worldwide such as Sinovac, Novavax. However we have not found any country using these vaccines and providing complete data. In the supplement we discuss some partial results from the incomplete data available from Mongolia (not the ratios), which seem to confirm the quality of these vaccines as compared to the others.

**Fig. 5.**
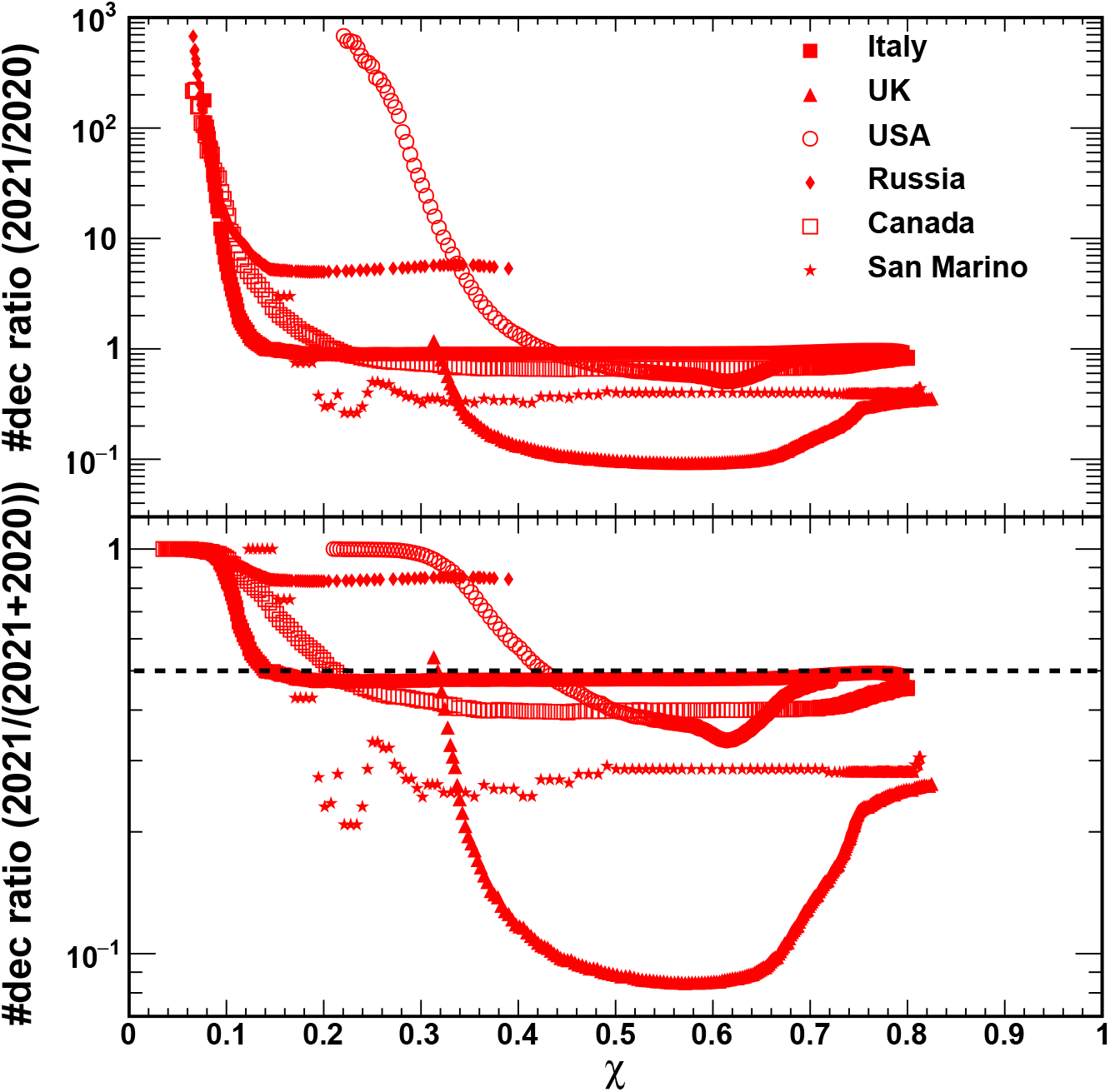
(Color online) (Top) Deceased case ratio (2021/2020) and (Bottom) Deceased case ratio (2021/(2020+2021)) of the same time period in 2021 and 2020 versus vaccination ratio *χ* for the countries indicated.

As we discussed above, the divergence seen at low *χ* is due to low statistics data at the beginning of the pandemics, i.e., early 2020. In order to compare different responses on the same scale we define the ratio of the period 2021/(2020+2021). In this case if the number of deaths in the period 2021 is much larger than the corresponding period in 2020, the ratio converges to 1. This is seen in figure 5 (bottom panel) for very low *χ* and it is due to statistics at the beginning of the pandemics. If the vaccines are not working, because of a new variant or loss of efficacity with time, then the ratio should converge to 0.5, dashed line in the figure 5 (bottom panel). The latter is observed for Italy, USA and Russia (larger than 0.5). Canada seems to be approaching the 0.5 value while the UK and RSM are well below it meaning that the vaccines used in those countries are still effective. We notice the increase of the ratio for the UK at larger vaccination rates, corresponding to later times, which may be due to the choice of the UK government to use other vaccines instead of the AstraZeneca (AZ) for people below 40 years of age. This was due to the claim that the AZ vaccines may provoke blood clots. This claim was largely amplified by the press leading to many countries to halt or slow down the use of this particular vaccines. In the UK, 49 blood clots cases have been reported after 28.5 million AZ vaccines have been administrated, out of about 49 million vaccines administrated in the same period (Pfizer is the other one). This is about 2 cases per million people which is about double the value observed normally in the population, thus a negligible factor. Nevertheless many countries opposed it or slowed down its use (Germany for instance). Clearly the economical factor is huge, recall that the AZ vaccine was led by the university of Oxford-UK with the intent of creating a vaccines readily available to all for a low price, about $2, i.e., an order of magnitude less than other more ‘popular’ vaccines such as Pfizer and Moderna, see table 2. Our results show a quite different scenario about the vaccines efficacity, thus while rich countries should continue their economical games, ‘third world’ countries can confidently rely on the AZ vaccine. One question remains is why prices are so much different.

**Table 2.** The prices of the vaccines, data are taken from ref. [9].

| Vaccine name | Type | Price (dollar/dose) | total doses |
| --- | --- | --- | --- |
| Pfizer-BioNTech | mRNA | 19.5 | 2 |
| Moderna | mRNA | 25-37 | 2 |
| AstraZeneca | Adenovirus-based | 2.15(EU), 3-4(UK), 5.25(South Africa) | 2 |
| Johnson & Johnson | Adenovirus-based | 10 | 1 |
| Russia's Sputnik V Vaccine | Adenovirus-based | 10 | 2 |
| Sinovac Biotech | Inactivated SARS-CoV-2 virus | 29.75 | 2 |
| Novavax | Protein-based | 16 | 2 |
| CanSino Biologics | Viral vector | unknown | 1 |
| Bharat Biotech | Inactivated SARS-CoV-2 virus | 2 | 2 |

A comparison between figures 4 and 5 confirms that herd immunity may be not reached anytime soon with the notable exception of the ratio of the number of deceased for the UK and RSM. This may suggest further vaccinations for the other countries and maybe we should consider a wider usage of the AstraZeneca vaccine as confirmed by the different observables used in this work.

## 3 Conclusions

In this work we have explored the similarities and differences of COVID-19 with the Spanish flu of 1918 using the logistic map as guidance. We have made some prediction on the number of cases and estimated the progress reached through vaccinations. We have shown that herd immunity cannot be reached because of the Delta variant. However, the death probability has been largely reduced and it affects almost exclusively the unvaccinated. A comparison of different countries using different vaccine types and different policies to contrast the pandemic shows a larger efficacity for the UK, Norway, S. Korea and other countries. Cases are bouncing back (especially the positives) to the values of the previous year thus suggesting a loss of vaccines efficacity and the possibility for a third dose. Countries like the UK and the Republic of San Marino seem still doing well thus may be reasonable to offer to the population a third dose with the AstraZeneca or the Sputnik-V vaccines, given a green light from the doctors for each case. Also, suitable blood test investigations for the vaccinated people should be performed before administrating the third dose to exclude people who have enough antibodies. A statistics of cases for different vaccine types can help decide which route to follow to produce new vaccines able to confront possible new and more deadly variants if any. At this stage it seems to us not useful to implement further actions to force people to get vaccinated if they do not want to do that. The most important action in those cases is information. People that gets positive to Covid have 1 probability out of 50 to die. Since about 70-80% of the positives are vaccinated and their probability to die is close to zero, it means that the unvaccinated positive to Covid have probability 1 out of 10-15 to die. Furthermore, given the high probabilities to be positive to the virus (about 20-30% of the tested), sooner or later we may become positive and it is better to be ready, i.e. vaccinated, to reduce the risk practically to zero to die.

## Supporting information

updated figures to dec.21,2021

## Data Availability

All data produced in the present study are available upon reasonable request to the authors

## Acknowledgments

This work was supported partly by the National Natural Science Foundation of China (Grant Nos. 11905120 and 11947416) and by the U.S. Department of Energy under Grant No. DE-FG03-93ER40773 and NNSA Grant No. DENA0003841 (CENTAUR).

“ Note added in proof: extending the analysis of this paper to December 21, 2021 did not show any relevant changes due to the Omicron variant. Updated figures are available upon request from the authors”.

## Author contributions

A.B. designed the research and drafted the work, H.Z. collected the data, conducted the data analysis and revised the draft.

## Supplemental material

In this supplement we discuss different countries, which adopted different strategies to contrast COVID-19. Sweden was one of the few countries letting the disease to spread freely and as a results had the largest number of fatalities as compared to nearby countries like Norway and Denmark, see figure S1. When divided by the population, their death ratio is a factor 10 larger than Norway shedding some bad light on the choice of herd immunization. Probably that is also the reason why the Sweden data is incomplete, see the bottom part of figure S1. This is the worst pandemic for Sweden since the S1918 [12]. S. Korea adopted the strategy of tracking, attention to hygiene and masks decreasing the fatality rate of a factor 2 respect to Norway. Thus there are better methods to contrast the virus even without vaccine and these are important lesson for future pandemics.

---

**Fig. S1.**
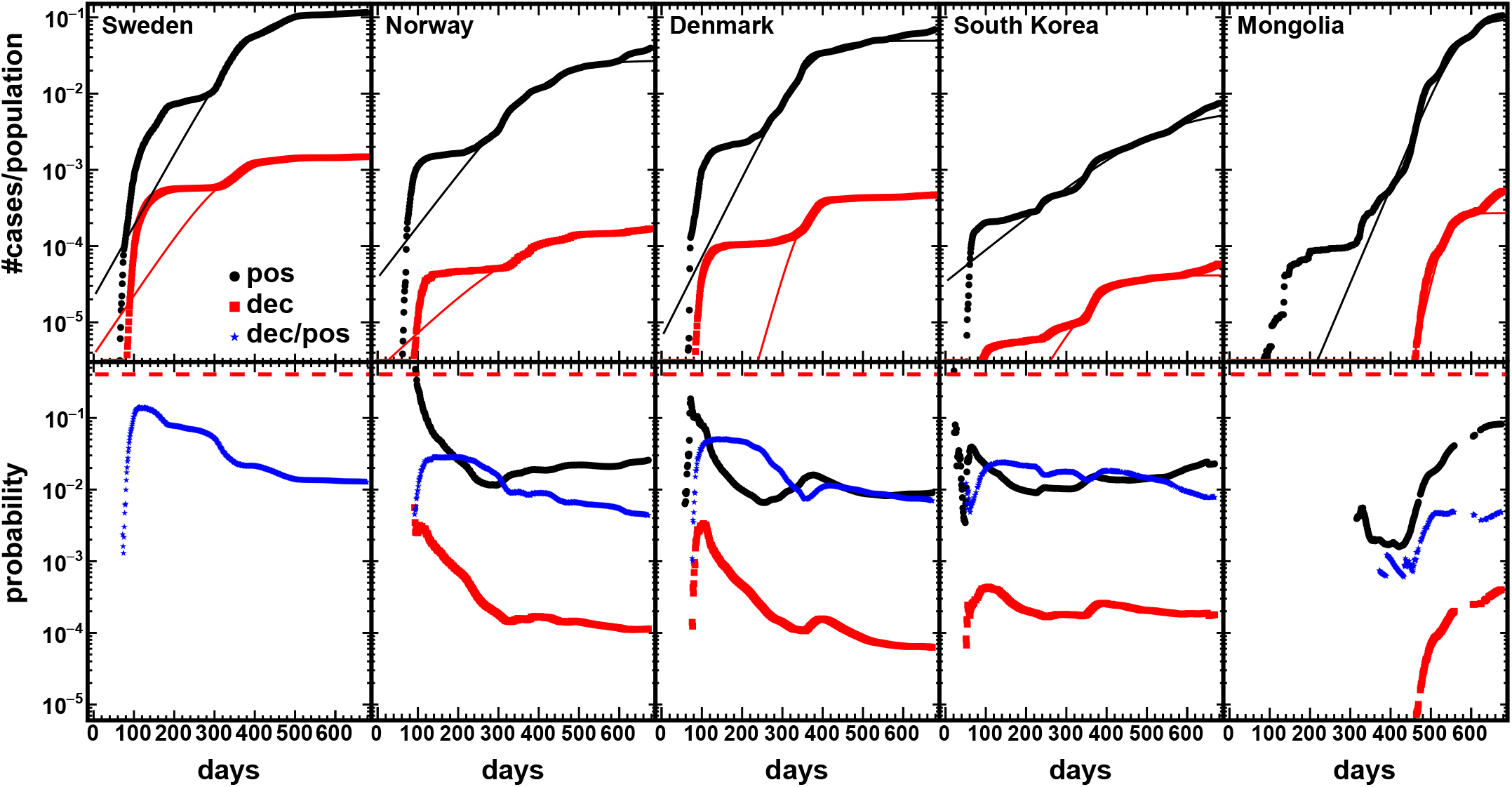
(Color online) Same as fig. 1 for different countries. Updated to November, 2021.

**Fig. S2.**
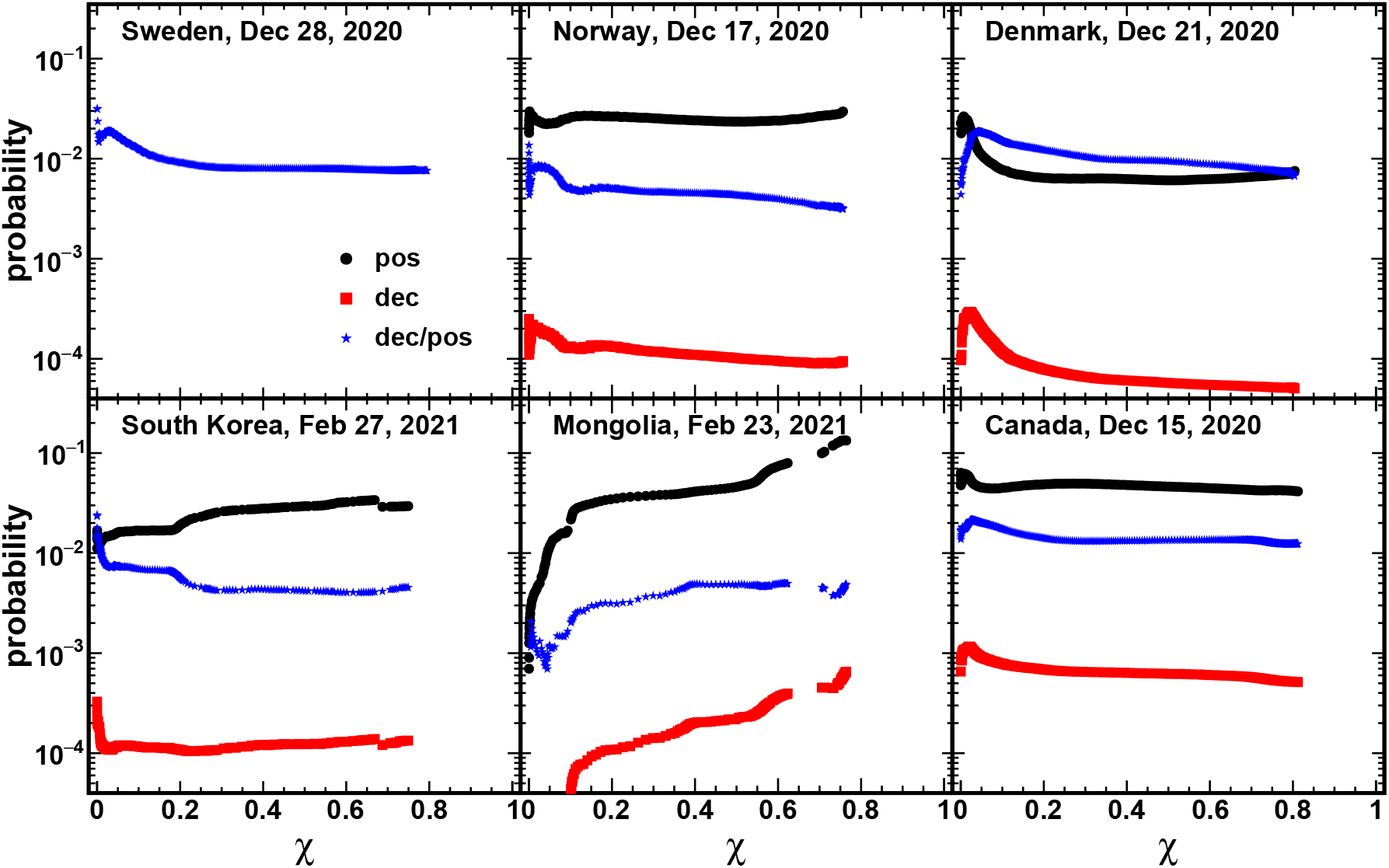
(Color online) The day 1 is the vaccination time which is shown in the legend. In figure S2 we plot the probabilities as function of vaccination rate for the same countries of fig. S1, compare to figure 3.

## References

1. Sabbatini, S. et al., “Le infezioni in medicina” n. 4, 272–285 (2007).

2. Turcotte, D. L., Fractals and chaos in geology and geophysics (Cambridge University Press, 1992).

3. Schuster, H. G., Deterministic Chaos (VCH, 1995).

4. Baran, V., Zus, M., Bonasera, A., Paturca, A., Rom. Journ. Phys. 60, 1263 (2015).

5. Bonasera, A., and Zhang, S., Front. Phys. 8, 171 (2020).

6. Bonasera, A., Bonasera, G. and Zhang, S., Eur. Phys. J. Plus 135, 453 (2020).

7. Zheng, H. and Bonasera, A., Eur. Phys. J. Plus 135, 799 (2020).

8. Zheng, H. and Bonasera, A., https://www.medrxiv.org/content/10.1101/2020.11.11.20229872v2 (2020)

9. https://www.biospace.com/article/comparing-covid-19-vaccines-pfizer-biontech-moderna-astrazeneca-oxford-j-and-j-russia-s-sputnik-v/

10. https://covid19.who.int/WHO-COVID-19-global-data.csv

11. https://ourworldindata.org/grapher/full-list-total-tests-for-covid-19

12. Ledberg, A., Frontiers in Public Health, Vol 9, 579948 (2021).

