## Supplementary figures and images for "Controlling the worldwide chaotic spreading of COVID-19 through vaccinations"

### appendix_figure1_3.pdf

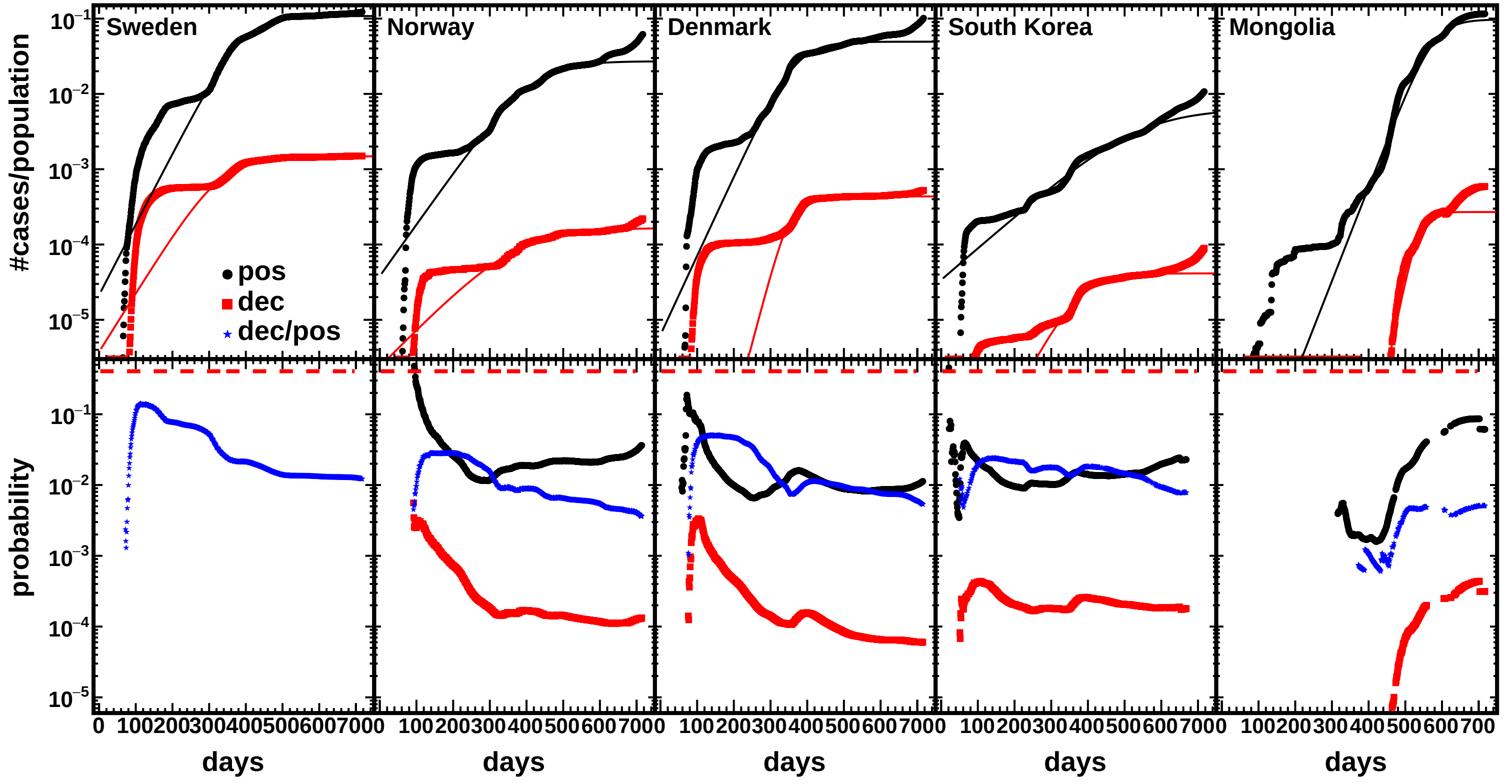

### dec_case_ratio_2panels.pdf

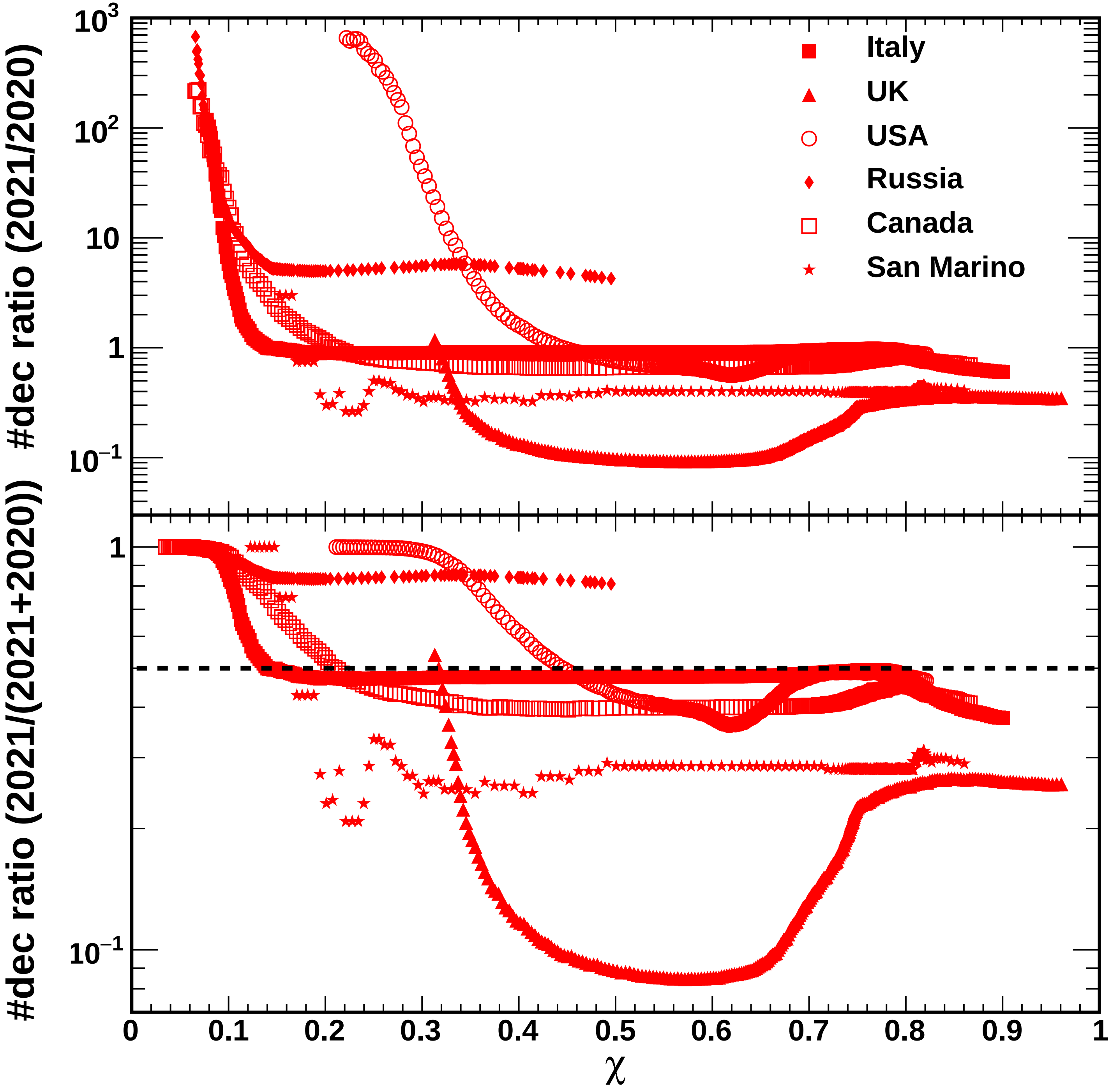

### f3_chi_prob.pdf

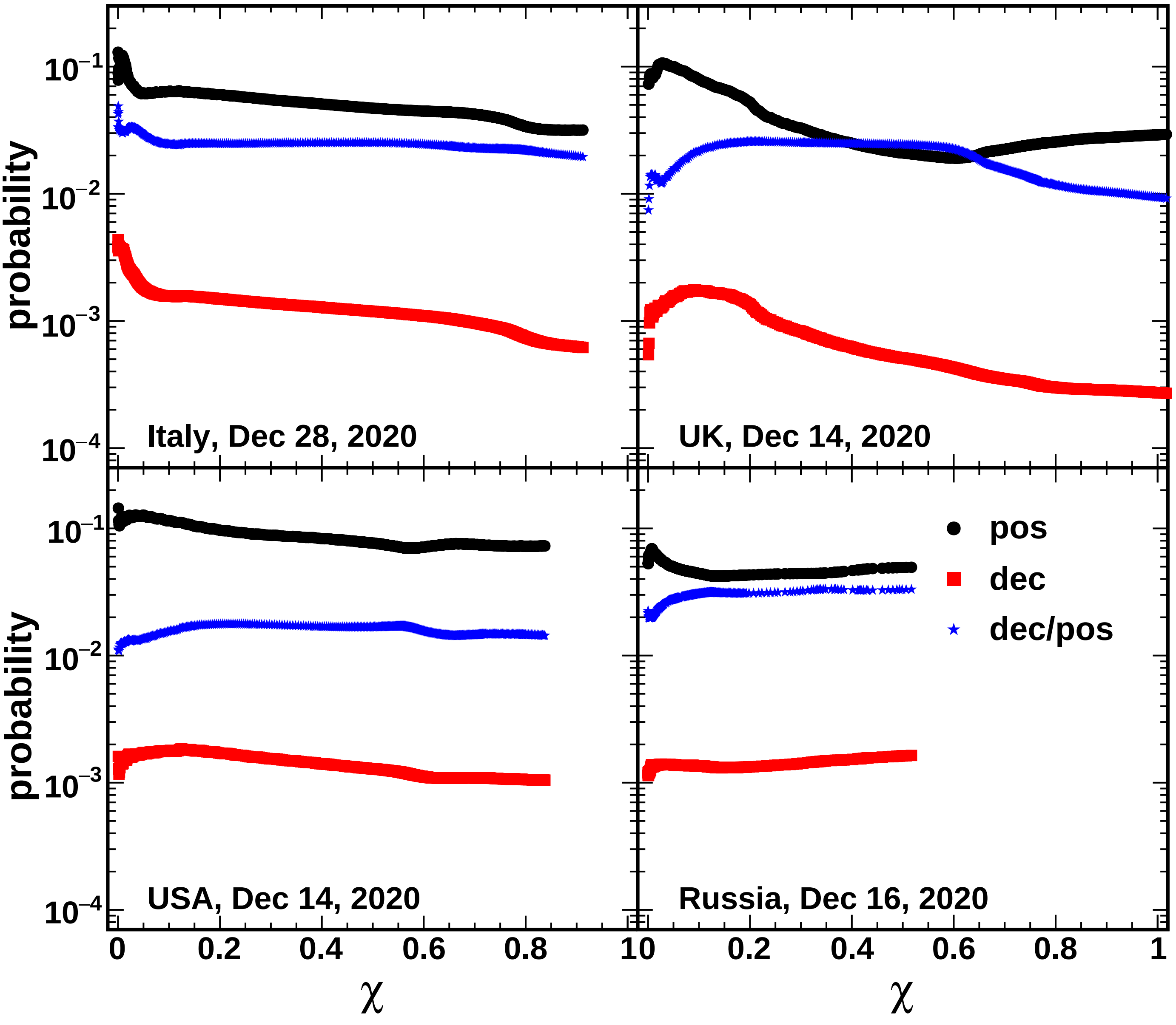

### figure_italy_uk_2.pdf

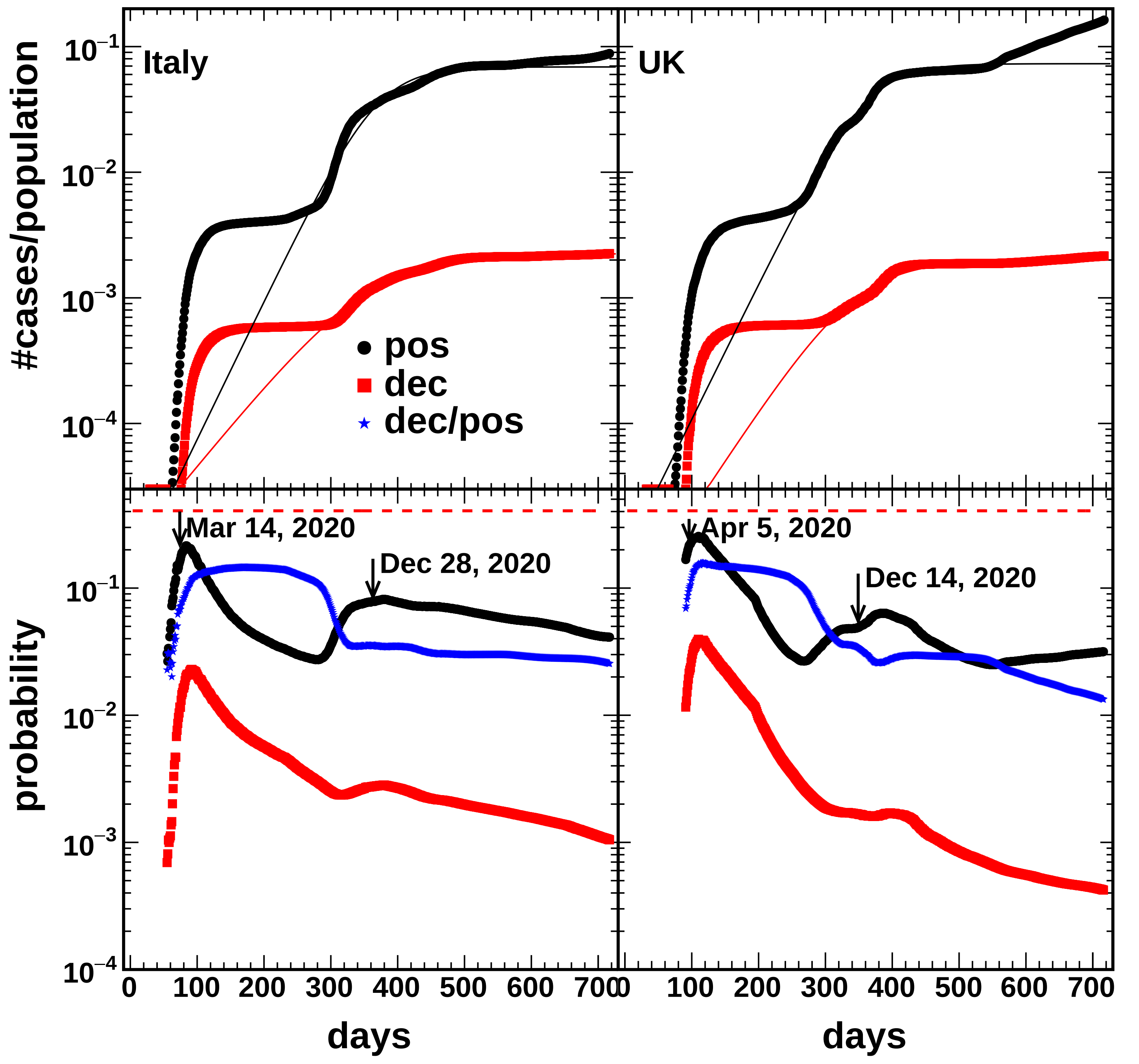

### it_uk_map.pdf

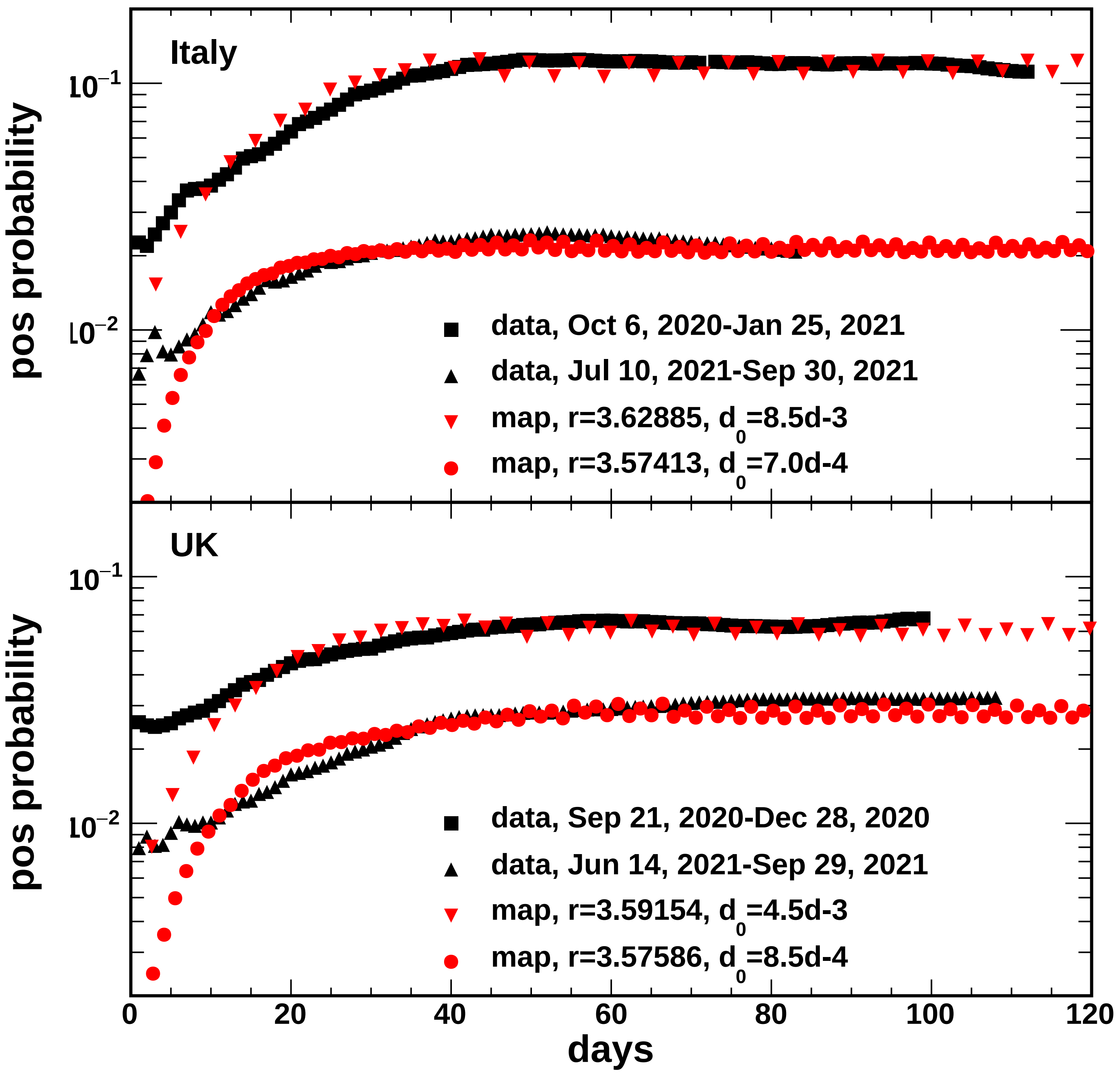

### prob_ratio_versus_chi.pdf

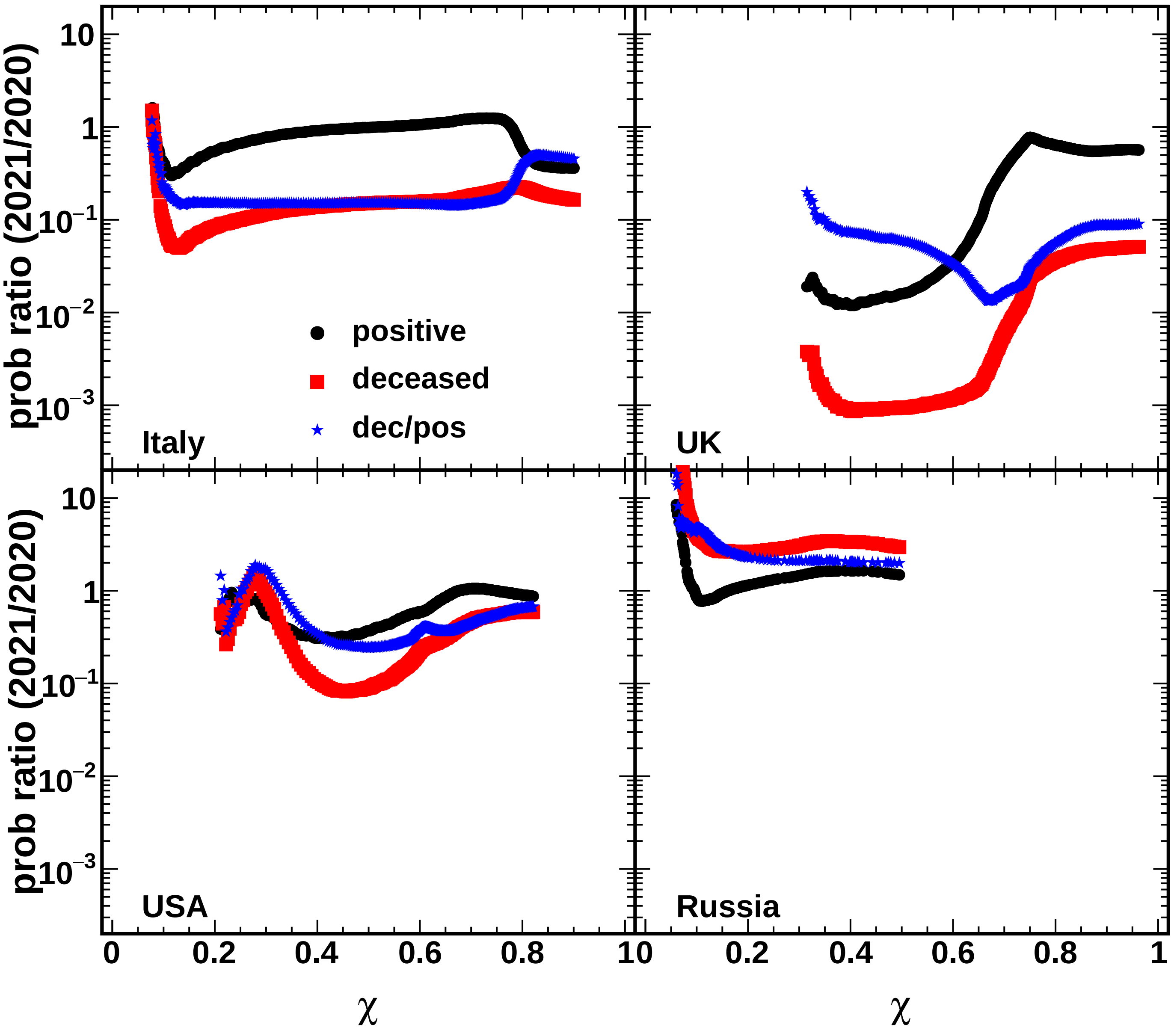

### s2_chi_prob.pdf

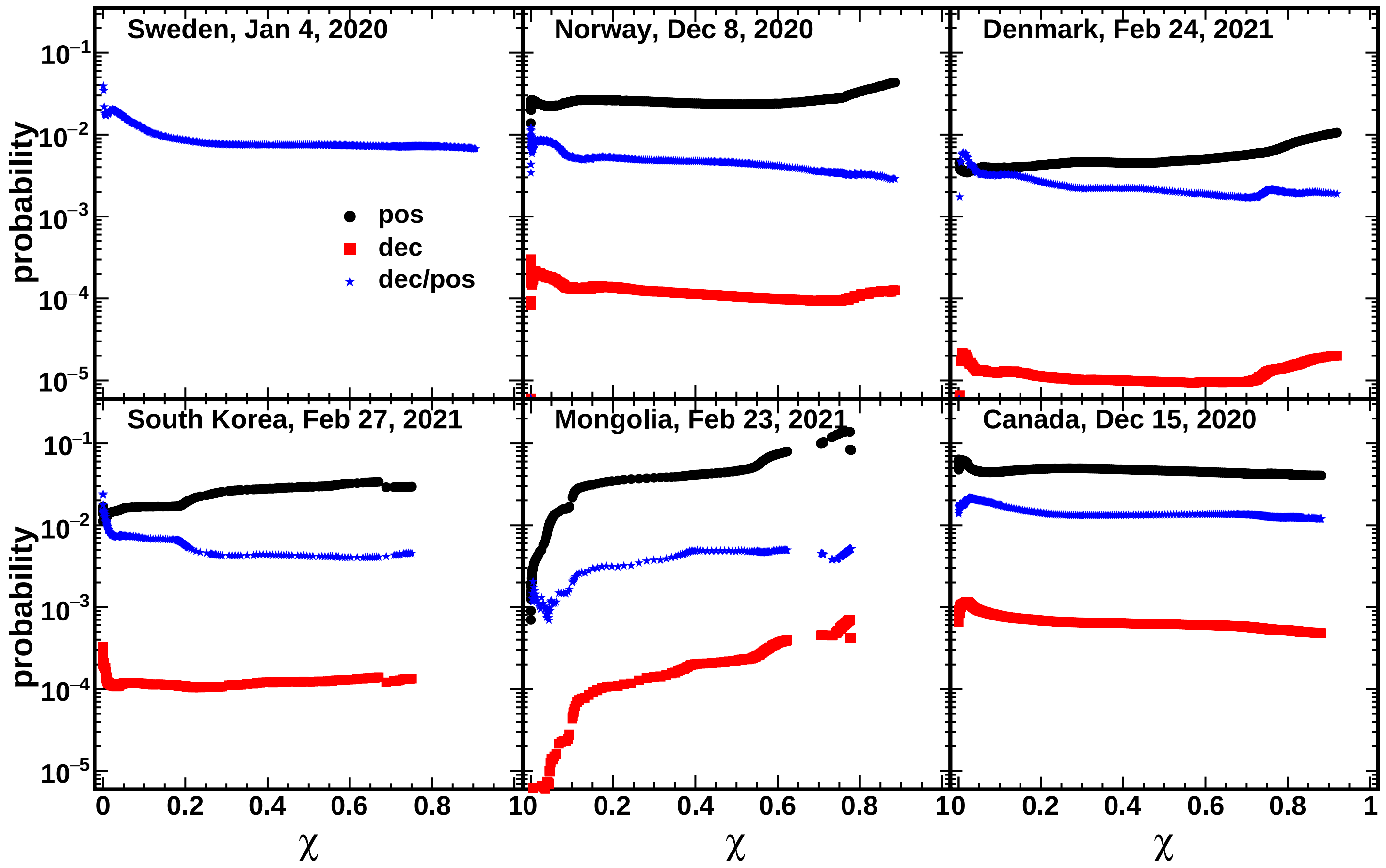
